# The association between likely ADHD and autism, and not being in employment, education, or training: a frequency-matched case-control study in working-age adults in the UK

**DOI:** 10.64898/2026.08.03.26359585

**Authors:** Lisa Quadt, Emma Russell, Jane Green, Emily Joynson, Beth Jones, Ulrich Müller-Sedgwick, Conor Davidson, Hugo Critchley, Jessica Eccles

## Abstract

**Background:** To estimate the frequency of likely, often undiagnosed autism and attention deficit hyperactivity disorder (ADHD) in UK adults of working age (18-66 years) who are not in education, employment, or training (NEET), and to examine whether neurodivergent traits are associated with NEET status directly and indirectly via health burden and educational attainment.

**Design:** Frequency-matched online case-control study.

**Setting:** UK general population, recruited via online research platform Prolific.

**Participants:** Six hundred adults of working age (18-66 years); 300 NEET, 300 in education, employment, or training (EET) matched at the marginal level on age, sex assigned at birth, and ethnicity.

**Primary and secondary outcome measures:** Autistic traits (Ritvo Autism and Asperger Diagnostic Scale–14, RAADS-14) and ADHD traits (Adult ADHD Self-Report Scale, ASRS-5) indexed likely autism and ADHD (cut-off ≥14 on each). Physical and mental health conditions were self-reported and aggregated into composite indices of health burden. NEET status was the primary outcome; educational attainment and health burden were tested as parallel mediators of the association between neurodivergent traits and NEET status.

**Results:** NEET participants screened positive more frequently for likely autism (66.3% vs 49.0%; OR 2.05, 95% CI 1.48 to 2.85) and likely ADHD (34.3% vs 26.0%; OR 1.46, 95% CI 1.03 to 2.08) than EET participants, despite identical existing formal diagnosis rates. Physical (OR 2.00, 95% CI 1.38 to 2.90) and mental (OR 2.57, 95% CI 1.80 to 3.68) health conditions were also associated with higher odds of NEET status. In mediation analyses, neurodivergent traits predicted NEET status both directly (OR 1.33, 95% CI 1.13 to 1.68) and indirectly via greater health burden (indirect OR 1.13, 95% CI 1.02 to 1.35) and lower educational attainment (indirect OR 1.08, 95% CI 1.03 to 1.15).

**Conclusion:** NEET adults showed a marked excess of autism and ADHD traits, alongside elevated physical and mental health burden and lower educational attainment. Earlier recognition of neurodivergent traits, proactive provision of equitable requirements in education and employment, and integrated physical and mental health support may reduce NEET risk in this population. This has considerable implications for policy and practice in health, education, and wider society.

## Introduction

Disabled people and those with long-term health conditions are substantially overrepresented among adults not in employment, education, or training (NEET), yet the precise contribution of neurodivergence (e.g., autism, attention deficit hyperactivity disorder; ADHD) to this disparity remains poorly characterised,^1^ ^2^ particularly in regard to their co-occurrence and their impact on health and education. This characterisation gap itself is partly due to how neurodivergent conditions are identified: diagnostic systems were not designed to recognise the full range of neurodivergent presentations,^3^ ^4^ and large numbers of autistic people and those with ADHD, particularly women, older adults, and people from the global majority, remain undiagnosed well into adulthood.^5–8^ Studies relying on formal diagnosis therefore likely report substantial underestimates of neurodivergence in NEET populations, leaving the true scale of this relationship obscured. Here, we address this gap and provide estimates of likely, undiagnosed neurodivergence in NEET adults.

Recent UK labour market data indicate that this challenge is large-scale and worsening: in January to March 2026, an estimated 1,012,000 young people aged 16-24 were NEET (13.5%), which marks an increase of 89,000 compared to the previous year and the first time this measure has exceeded one million in over a decade.^9^ Concurrent declines in job vacancies alongside rising competition for available roles further suggest tightening labour market conditions that are likely to disproportionately affect individuals with health-related barriers to participation.^10^ Capacity for assessments for ADHD and autism, and post-diagnostic support for these conditions far outstrip demand (reference), and represent a significant unmet need.

These trends have prompted renewed policy attention. The recent independent Milburn Review of Young People and Work characterises the current rise in youth disengagement as a “whole-system failure”, driven by interaction between labour markets, education systems, welfare provision, and health.^10^ The review reflects not only individual vulnerability but a persistent mismatch between institutional structures and diverse needs.

Within this context, neurodivergence likely plays a substantial yet insufficiently characterized role. Autism and ADHD are associated with differences in executive functioning, social communication, and emotional regulation that can affect educational attainment and labour market participation, especially when they remain unidentified and/or unsupported.^11^ ^12^ Longitudinal cohort studies indicate that both conditions are independently associated with increased likelihood of NEET status in early adulthood, and population-based analyses consistently show lower employment rates among autistic adults and individuals with ADHD compared with the general population.^1^ Despite this, estimates of neurodivergence in NEET populations rely primarily on formal diagnoses, which are prone to underestimate true prevalence due to diagnostic biases.^13^

Structural features of standard educational and occupational settings, including sensory demands, rigid organisational structures, and normative expectations of social functioning present barriers for some individuals.^14^ ^15^ Neurodivergent people are also more likely to experience physical and mental health problems,^16^ creating an additional barrier to participation in education and employment. Importantly, under UK equality legislation, entitlement to equitable requirements (often referred to as reasonable adjustments)^17^ is based on “functional impairment” rather than diagnostic status; however, recognition of neurodivergence is often a necessary step to understanding what kind of support will enable and improve participation in education and work. Furthermore, access to accommodations are often facilitated by formal diagnosis,^18^ and evidence suggests that workplace adjustments, supported employment programmes, and inclusive educational environments can improve participation outcomes among neurodivergent individuals.^19^

In this study, we examined the prevalence of autistic and ADHD traits in a frequency-matched sample of NEET and non-NEET (in education, employment, or training; EET) adults in the UK. We further investigated differences in mental and physical health and explored perceived environmental enablement in educational and occupational contexts.

## Methods

### Study design, setting, and participants

In this frequency-matched case-control online study, participants were recruited via the platform Prolific and completed an online survey on the platform Qualtrics. We recruited 600 adult participants (n=300 per group). Eligible participants were adult UK residents between the ages of 18-66 years who were fluent in English and not retired. In the NEET group, eligible participants were currently not in employment, education, or training. In the EET group, eligible participants were currently in employment, education, or training.

Participants in the EET group were frequency matched at the marginal level on age, sex, and ethnicity to the NEET group. Target frequencies were derived from the NEET sample, and independent demographic quotas were implemented in Qualtrics. Recruitment of EET participants continued until the target marginal distributions were met. Participants who entered the survey after a demographic quota had been filled were screened out.

This study was approved by the Brighton and Sussex Medical School Faculty Research Ethics Committee (Reference #2026-1435-1841).

### Measures

#### Demographic, employment, and socio-economic information

We asked participants to provide information on their age, sex assigned at birth, ethnicity, and highest level of education. NEET participants were asked for the main reason they are not in employment, education, or training, the length of their NEET status, main source of income, reason for leaving employment, education, or training (if applicable), and whether they wish to enter employment, education, or training in the future. EET participants were asked for their current EET status, whether they received income support from the government, and annual income.

We also asked both groups about accessibility factors in employment and education contexts; in the NEET group, based on Grant et al (2026)^15^ we asked to what extent their previous place of employment, education, or training (if applicable) enabled them to control their sensory environment, manage any specific conditions they may have, and express themselves authentically, and how important these aspects would be in a future place of employment, education, or training. For the first set of questions, participants answered on a 5-point Likert scale ranging from “Not at all” (1) to “A great deal” (5). For the second set of questions, participants answered on a 5-point Likert scale ranging from “Not at all important” (1) to “Extremely important” (5). EET participants were asked the same questions with the same answer options about their current workplace/education environment (i.e., how important these factors are to them and to what extent they are actually realized in their current working/education environment).

#### General health information

All participants were asked to indicate whether they had physical and/or mental health conditions, and to list these conditions if applicable.

Self-reported physical and mental health conditions were provided as free-text responses and categorised using a predefined, rule-based framework that was manually reviewed for accuracy. Physical conditions were grouped into 12 categories of conditions (sensory, neurological, pain/musculoskeletal, cardiovascular, respiratory, endocrine/metabolic, gastrointestinal, autoimmune, gynaecological/reproductive, cancer, fatigue, mobility), and mental health conditions into nine categories of conditions (depressive disorders, anxiety disorders, obsessive compulsive disorder, trauma disorders, bipolar disorders, personality disorders, eating disorders/additions, other).

Within each category, count variables were created reflecting the number of distinct conditions reported (e.g. individuals reporting both generalised anxiety disorder and panic disorder received a score of 2 in the anxiety disorders category). Composite variables representing the total number of physical health conditions, mental health conditions, and a combined overall count were created to operationalise physical, mental, and overall health burden.

#### Neurodivergence

All participants were asked to complete the six-item Adult Self-report ADHD Scale (ASRS-5)^20^ to index ADHD traits, and the 14-item Ritvo Aspergers and Autism Diagnostic Scale (RAADS-14)^21^ to index autistic traits. These are common screening questionnaires to assess whether an individual should be considered for a full diagnostic assessment if they meet screening threshold (score ≥ 14 for each).

We derived a composite “neurodivergent traits” measure by averaging the z-standardized total scores from the ASRS-5 and RAADS-14.

### Sample size

Formal sample size calculations were conducted for a two-group comparison of proportions (NEET vs non-NEET), using a two-sided test with a 5% significance level and 90% power. Based on existing evidence indicating that approximately 4–5% of adults screen positive for ADHD on the ASRS-5,^22^ and policy data suggesting substantially lower employment rates among neurodivergent adults,^23^ we anticipated a meaningful difference in ADHD screening prevalence between groups. To ensure sufficient power to detect moderate effects and smaller, policy-relevant differences, we based calculations on a conservative minimum detectable effect size of Cohen’s ℎ ≈ 0.26. This corresponds to an absolute difference in screening prevalence of approximately 6–8 percentage points (e.g., 5% vs 11–13%). Under these assumptions, a sample size of approximately 300 participants per group was required.

## Statistical analyses

Analyses were conducted in IBM SPSS Statistics Version 30. Group characteristics and statements on accessibility factors were summarised using descriptive statistics. Between-group differences in categorical variables were examined using chi-square tests; between-group differences in continuous variables were examined using independent-samples t-tests or Mann-Whitney U tests.

Binary logistic regression was used to estimate odds ratios (ORs) with 95% confidence intervals (CIs) for the association between NEET status and (i) likely autism (RAADS-14 score ≥14) and (ii) likely ADHD (ASRS-5 score ≥14), entered as separate dependent variables. Each model was run unadjusted and subsequently adjusted for educational attainment. An equivalent approach was applied for physical health status and mental health status. Model fit was assessed using the omnibus chi-square test, Nagelkerke R², and overall classification accuracy.

We divided NEET and EET groups by likely neurodivergence status (if both or one of RAADS-14 or ASRS-5 were above threshold) to compare accessibility factors within groups using independent samples t-tests.

Parallel mediation analysis was conducted using SPSS PROCESS macro (Model 4) with 5,000 bootstrap samples to examine whether overall health burden and educational attainment mediated the association between neurodivergent traits and NEET status. Neurodivergent traits were specified as the independent variable, NEET status as the binary outcome, and health burden and educational attainment as simultaneous mediators. Logistic regression models were estimated for all paths, and effects are reported as log-odds and odds ratios (ORs) with 95% bias-corrected confidence intervals (CIs).

## Results

### Participants

All demographic and clinical group comparisons are displayed in Table 1. Level of education differed significantly between groups and was included as a covariate in subsequent analyses.

**Table 1:** Demographic and clinical group comparisons.

| Baseline characteristic | NEET | EET | Test-statistic (df) | p-value |
| --- | --- | --- | --- | --- |
| **Age (mean (SD))** | 37.5 (11.5) | 38.2 (11.5) | $t(598) = .726$ | .234 |
| **Sex assigned at birth** | | | $\chi^2(2) = 1.00$ | .606 |
| Female | 149 (49.7%) | 149 (49.7%) |  |  |
| Male | 150 (50%) | 151 (50.3%) |  |  |
| Intersex | 1 (0.3%) | - |  |  |
| **Ethnicity** | | | $\chi^2(3) = 0.066$ | .996 |
| Asian or Asian British | 29 (9.7%) | 28 (9.3%) |  |  |
| Black, Black British, Caribbean or African | 13 (4.3%) | 13 (4.3%) |  |  |
| Mixed or multiple ethnic groups | 13 (4.3%) | 12 (4%) |  |  |
| White | 245 (81.7%) | 247 (82.3%) |  |  |
| **Education <sup>a</sup>** | | | $\chi^2(4) = 49.98$ | <.001 |
| Attended college, no degree | 36 (12%) | 37 (12.3%) |  |  |
| A-levels or similar | 59 (19.7%) | 40 (13.3%) |  |  |
| GCSE or similar | 78 (26%) | 23 (7.7%) |  |  |
| A-levels or similar | 59 (19.7%) | 40 (13.3%) |  |  |
| Graduate degree | 47 (15.7%) | 77 (25.7%) |  |  |
| **Physical health conditions** | | | $\chi^2(1) = 15.57$ | <.001 |
| Yes | 119 (39.7%) | 73 (24.3%) |  |  |
| Physical health burden (mean (SD)) | 0.59 (1.00) | 0.37 (1.96) | $U = 37,953$ | <.001 |
| **Mental health conditions** | | | $\chi^2(1) = 32.61$ | <.001 |
| Yes | 149 (49.7%) | 80 (26.7%) |  |  |
| Mental health burden (mean (SD)) | 0.74 (1.20) | 0.37 (0.75) | $U = 33,995.5$ | <.001 |
| **Overall health burden (mean (SD))** | 1.47 (1.78) | 0.74 (2.27) | $U = 31,370$ | <.001 |
| **Existing Diagnoses** |  |  |  |  |
| Autism <sup>b</sup> | 23 (7.7%) | 20 (6.7%) | $\chi^2(1) = 0.19$ | .659 |
| ADHD <sup>b</sup> | 23 (7.7%) | 20 (6.7%) | $\chi^2(1) = 0.19$ | .659 |
| **Likely Neurodivergence** |  |  |  |  |
| Likely Autism <sup>c</sup> | 199 (66.3%) | 147 (49%) | $\chi^2(1) = 18.46$ | **<.001** |
| Likely ADHD <sup>d</sup> | 103 (34.3%) | 78 (26%) | $\chi^2(1) = 4.54$ | **.033** |
| Likely AuDHD <sup>e</sup> | 95 (31.7%) | 66 (22%) | $\chi^2(1) = 7.14$ | **.008** |
| RAADS-14 Score (mean (SD)) | 19.5 (11.3) | 15.1 (10.7) | $t(598) = -4.85$ | **<.001** |
| ASRS-5 Score (mean (SD)) | 11.6 (4.8) | 10.21 (4.9) | $t(598) = -3.60$ | **<.001** |

Data are n (%) unless otherwise specified. <sup>a</sup>Based on UK educational system. <sup>b</sup>Self-reported existing diagnoses by a healthcare professional. <sup>c</sup>Indication of autism for scores on RAADS ≥ 14. <sup>d</sup>Indication of ADHD for scores on ASRS-5 ≥ 14. <sup>e</sup>AuDHD=Screen positive for both autism and ADHD. RAADS=Ritvo Asperger and Autism Rating Scale. ASRS=Adult ADHD Self-Report Scale.

Most participants in the NEET group (282, 94%) had previously been employed, in education, or training, and two thirds (226, 75.3%) replied “Probably yes” or “Definitely yes” when asked if they wish to be employed, in education, or training in the future.

In the EET group, most participants were working full-or part-time (261, 87%), 29 (9.3%) were self-employed, six (2%) were in education or training, and five replied “other”. The majority of the group were not receiving income support from the government (273, 91%). Twelve participants did not disclose their income bracket, and 171 (59.4%) earned above £30,000 per year.

### Likely neurodivergence and NEET status

Although the number of existing formal diagnoses of autism and ADHD did not differ between groups (Table 1), participants in the NEET group exceeded the screening threshold for likely autism and likely ADHD significantly more often. Meeting the RAADS-14 cut-off for probable autism was associated with approximately twofold higher odds of NEET status (OR 2.05, 95% CI 1.48–2.85, *p*<0.001), while meeting the ASRS-5 cut-off for probable ADHD was associated with around 50% increased odds (OR 1.46, 95% CI 1.03–2.08, *p*=0.033).

After adjustment for highest level of education, these associations remained statistically significant. The adjusted odds of NEET status were 1.85 (95% CI 1.32–2.60, *p*<0.001) for likely autism and 1.46 (95% CI 1.01–2.10, *p*=0.044) for likely ADHD. In both models, higher educational attainment independently predicted reduced odds of NEET status, with Ors of 0.67 (95% CI 0.59–0.76) in the autism model and 0.66 (95% CI 0.58–0.75) in the ADHD model (both *p*<0.001).

In terms of model performance, single-predictor neurodivergence models explained a small proportion of variance in NEET status (Nagelkerke *R²*=0.041 for autism; 0.010 for ADHD) and correctly classified 58.7% and 54.0% of participants, respectively. Adding education increased explained variance to 0.127 and 0.110 and improved classification accuracy to 62.3% and 62.2%, respectively, indicating modest but meaningful gains in model fit.

### Health and NEET status

The NEET group showed a statistically significant increased health burden (Table 1). Both physical and mental health conditions were also associated with increased odds of NEET status. In unadjusted models, reporting at least one physical health condition was associated with approximately double the odds of being NEET (OR 2.05, 95% CI 1.44–2.93, *p*<0.001), and reporting at least one mental health condition was associated with nearly threefold higher odds (OR 2.83, 95% CI 2.01–4.00, *p*<0.001).

These associations persisted after adjustment for educational attainment. In the adjusted models, having one or more physical health conditions remained associated with almost twofold higher odds of NEET status (OR 2.00, 95% CI 1.38-2.90, *p*<0.001), and having one or more mental health conditions remained associated with more than doubled odds (OR 2.57, 95% CI 1.80–3.68, *p*<0.001). As in the neurodivergence models, higher education consistently predicted lower odds of NEET (Ors 0.66–0.68, both *p*<0.001).

Health-only models explained 3–7% of the variance in NEET status (Nagelkerke *R²*=0.035 for physical health; 0.079 for mental health) and correctly classified 57.4% and 61.8% of cases, respectively. Adding education increased explained variance to 13-16% and improved overall classification accuracy to 63.1% (physical health+education) and 66.2% (mental health+education), yielding the highest explained variance across models.

### Parallel mediation analysis

Neurodivergent traits significantly predicted both greater health burden (*b*=0.54, OR = 1.71, 95% CI 1.43-2.05, *p*<.001) and lower educational attainment (*b*=−0.192, OR=0.83, 95% CI 0.73, 0.94, *p*=.003). In the outcome model, neurodivergent traits (OR=1.33, 95% CI 1.13-1.68, *p*=.002), health burden (OR=1.25, 95% CI 1.10, 1.44, *p*=.001), and educational attainment (OR=0.68, 95% CI 0.60-0.77, *p*<.001) each independently predicted NEET status (Nagelkerke *R²*=.162).

Both indirect pathways were significant: via health burden (OR=1.13, 95% CI 1.02-1.35) and via educational attainment (OR=1.08, 95% CI 1.03-1.15). The total indirect effect was also significant (OR=1.22, 95% CI 1.08-1.47). The direct effect of neurodivergent traits on NEET status remained significant (OR=1.33, 95% CI 1.13, 1.68, *p*=.002), indicating that overall health burden and educational attainment both contribute to the association between neurodivergent traits and NEET status (Figure 1).

**Figure 1:**
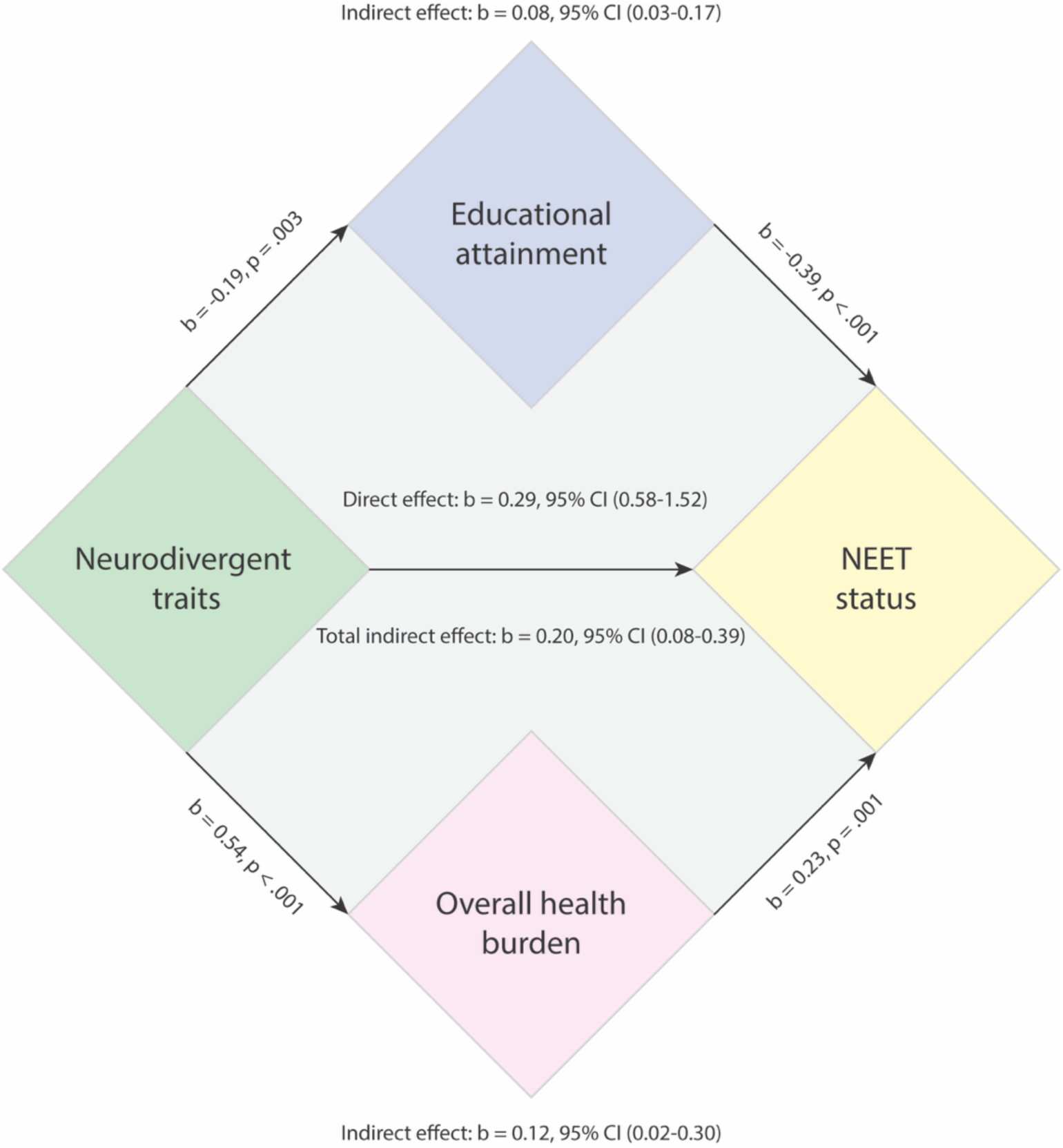
Parallel mediation model. Unstandardised path coefficients (b) are on the log-odds scale.

### Accessibility factors

Figure 2 displays response patterns in the NEET and EET groups about the extent to which their current (EET) or previous (NEET) workplace enables or enabled these factors, and how important these factors are to participants. While we did not statistically compare these between NEET/EET groups because they differ in what they refer to (actual current workplace/fictional future workplace), patterns can be observed. Most NEET participants answered “Not at all” or “A little” when asked about whether these accessibility factors were enabled in their previous place of work, while approximately two thirds of EET participants saw these enabled “A great deal” to “A moderate amount”. More than half of NEET participants found these factors to be “Extremely important” or “Very important” in a future place of work/education, while only around 40% of EET participants deemed them so.

**Figure 2:**
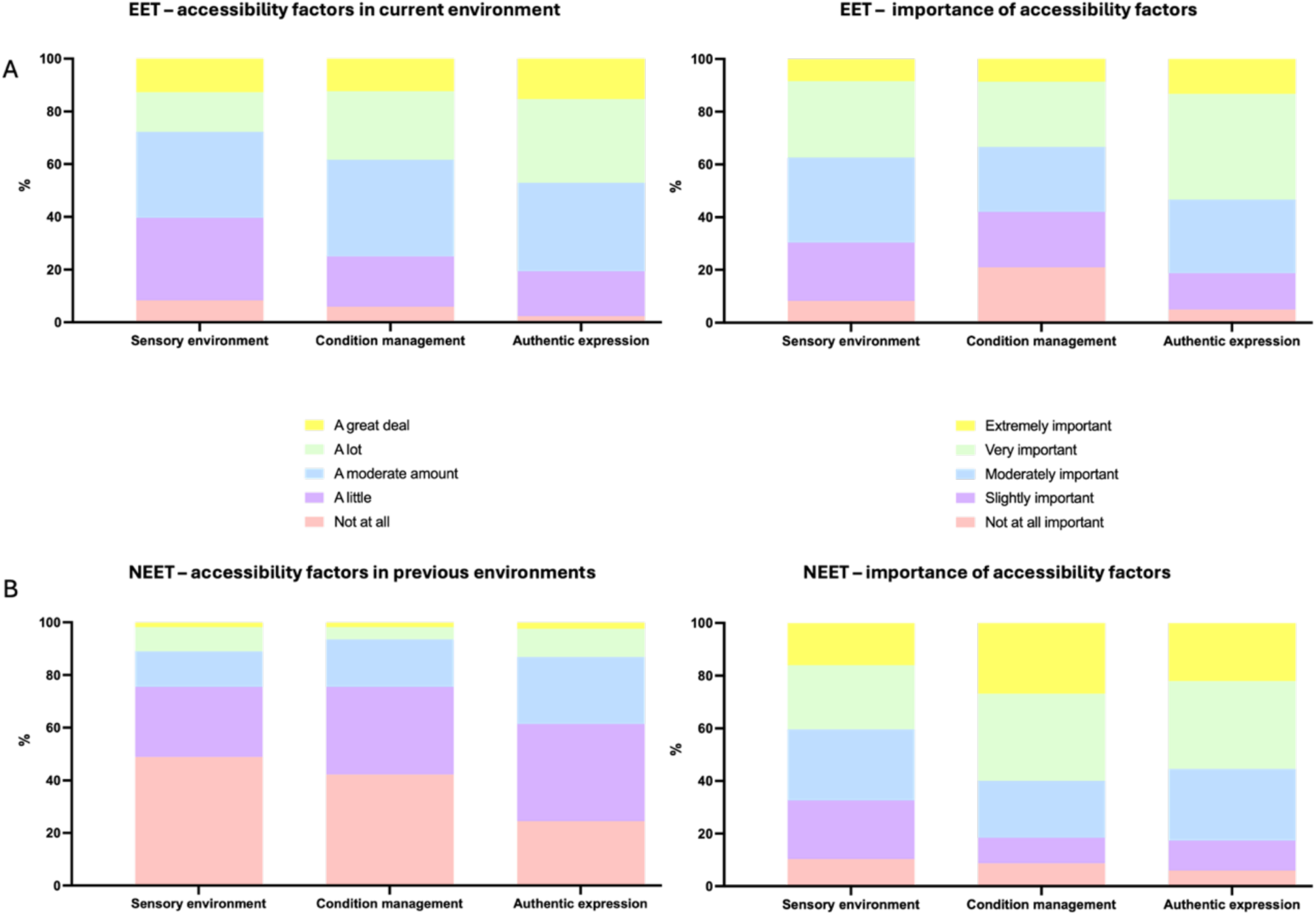
Accessibility factors in EET (A) and NEET (B) groups.

When stratified by likely neurodivergence status, broadly similar patterns were observed within EET and NEET groups (Figure 3). Across both groups, the clearest difference in experienced enablement related to authentic self-expression, which differed significantly by likely neurodivergence status in current EET environments (*t*(298)=3.13, *p*=.002; Fig. 3A) and previous NEET environments (*t*(280)=4.21, *p*<.001; Fig. 3C). By contrast, experienced sensory accessibility and condition management did not differ significantly within either group. Also in both groups, likely neurodivergent participant rated control over their sensory environment and management of any conditions as more important than those not classified as likely neurodivergent, both in current EET settings (*t*(298)=-5.58, *p*<.001; *t*(298)=-4.69, *p*<.001; Fig. 3B) and future NEET settings (*t*(298)=-5.95, *p*<.001; *t*(298)=-4.96, *p*<.001; Fig. 3D).

**Figure 3:**
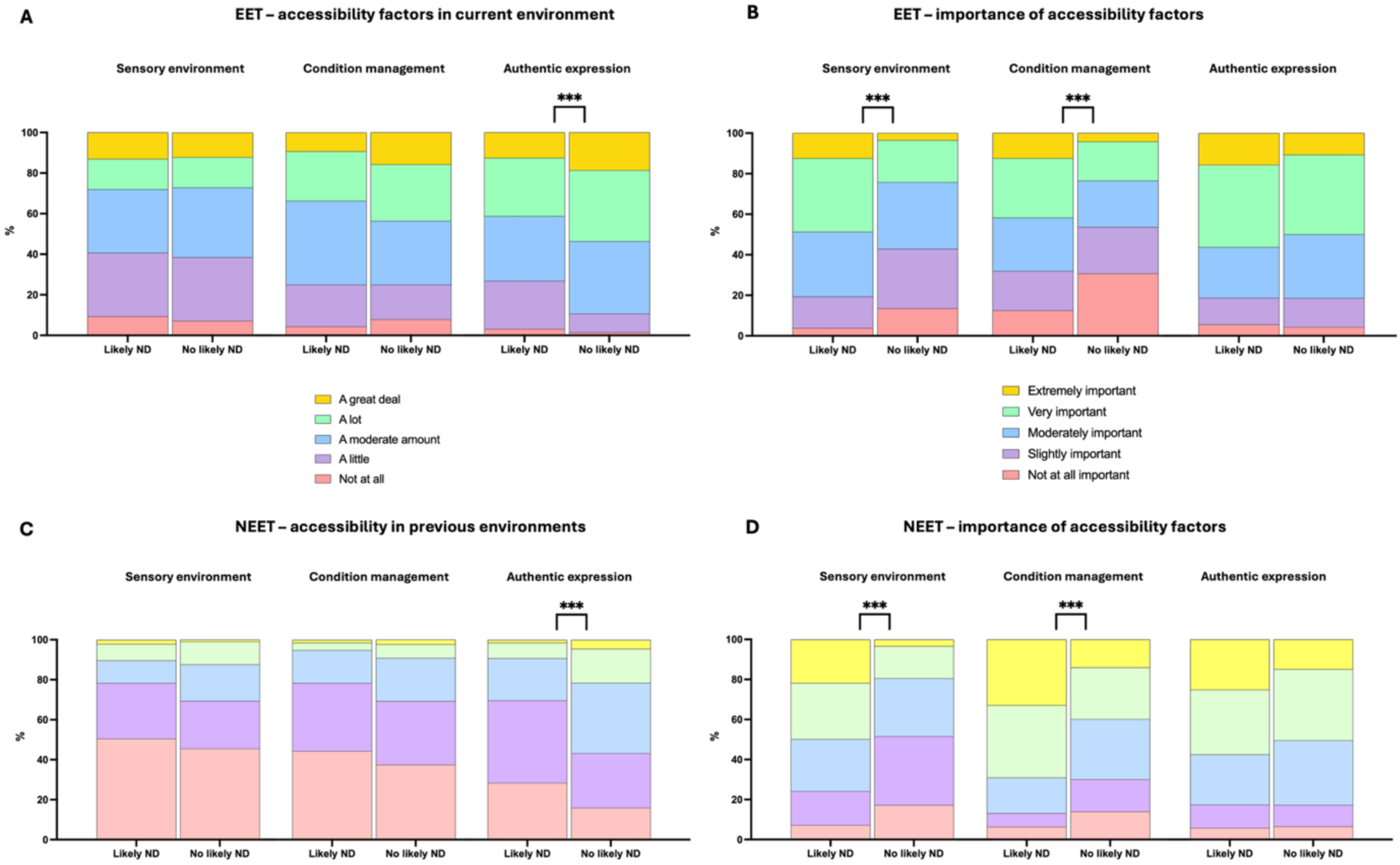
Accessibility factors in EET (A, B) and NEET (C, D) groups stratified by likely neurodivergence status.

## Discussion

This frequency-matched case-control study established that two-thirds of NEET participants screened positive for likely autism, and one-third screened positive for likely ADHD, yet formal diagnosis rates were identical across groups at 7.7% for each group. The eight-fold divergence between screening-positive and formally diagnosed rates in the NEET group is indicative of the unmet need for comprehensive diagnostic assessment and support for ADHD and autism, and associated conditions. Investing in this area has the potential to transform social outcomes and capital through their influence on employment, education, and training.

These findings replicate and extend longitudinal evidence from the ALSPAC cohort demonstrating that both autism and ADHD independently increase the odds of NEET status at age 25,^1^ and UK Biobank-derived data showing that neurodivergent adults are disproportionately concentrated in an employment–mental health trajectory characterised by economic inactivity and poor wellbeing.^24^

The absence of group differences in formal diagnosis rates, despite markedly elevated screening-positive rates in the NEET group, has direct clinical and policy implications. NHS diagnostic services are operating well beyond capacity: in 2023, only one in five individuals referred for autism or ADHD assessment was seen within target waiting times.^5^ Large numbers of neurodivergent adults are therefore more likely to enter, and remain in, NEET status without the identification or support that might enable the re-engagement that most NEET participants wished for their future.

Lower educational attainment was a significant independent predictor of NEET status in this sample, consistent with established evidence that autistic individuals without degree-level qualifications face substantially higher risk of unemployment,^25^ and that adults with ADHD are approximately four times more likely to occupy a peripheral labour market position.^26^ These data should not be interpreted as reflecting a fixed capacity deficit of neurodivergent people: lower attainment in this population is itself indicative of unmet support needs. Both autism and ADHD are associated with elevated rates of school dropout and qualification non-attainment.^1^ ^27^ Mechanisms that drive early disengagement from education are unsupported sensory, executive, and social-communication differences, combined with inadequate reasonable adjustments;^28–30^ these are the same mechanisms that subsequently entrench NEET status in adulthood. Addressing NEET vulnerability at its source therefore requires intervention in secondary education, not solely in employment services.

NEET status in this sample was not explained by neurodivergent traits alone. The mediation model showed that educational attainment and overall health burden are potential mechanisms that increase the likelihood of NEET status. This profile aligns with evidence that long-term physical health conditions substantially increase NEET risk,^31^ ^32^ and that persistent mental health difficulties arising in adolescence disrupt the school-to-employment transition.^33^ ^34^ Autistic adults and those with ADHD face elevated rates of co-occurring complex chronic conditions, such as chronic pain and chronic fatigue conditions^35^ (including Long COVID),^36^ hypermobility spectrum disorders,^37^ ^38^ depression and anxiety,^39^ ^40^ and bipolar disorder,^41^ intersecting vulnerabilities that are rarely captured together in either NEET research or neurodevelopmental and wider clinical practice. The present study addresses this gap directly, and its findings indicate that effective NEET intervention must address physical and mental health burden alongside neurodivergent need, rather than treating these as separate service domains. This requires upskilling the medical and educational workforce.

The perceived environmental enablement data add a practically important dimension. Stratifying by likely neurodivergence status showed that, in both NEET and EET participants, those screening positive attached greater importance to sensory control and condition management, and that authentic self-expression was the clearest domain where accessibility was reduced. NEET participants reported systematically poorer environmental enablement in previous educational or employment settings, and rated sensory control, the ability to manage their condition(s), and authentic self-expression as significantly more important in future settings than EET participants. These findings show that often, NEET status is not a reflection of individual deficit, but of a mismatch between neurodivergent need and environmental provision. The within-group analyses suggest that this pattern is not specific to NEET status. In both NEET and EET participants, likely neurodivergent individuals attached greater importance to sensory control and condition management, while authentic self-expression showed the clearest difference in experienced enablement. Together, these findings reinforce the interpretation that unmet accessibility needs, particularly around psychological safety and authentic self-expression, may represent a broader mechanism through which neurodivergent traits contribute to exclusion from education and employment. Reasonable adjustments, as mandated under the UK Equality Act 2010, are both the legal and the evidenced response to this mismatch; these data identify the specific adjustment domains; sensory environment, condition management, and psychological safety; that neurodivergent NEET adults themselves rate as most consequential for re-engagement.^15^ Employers and educational institutions that invest in these areas are not merely fulfilling legal obligations; they are addressing the modifiable structural factors that drive NEET status in this population.

### Strengths and limitations

Strengths of this study include the use of a frequency-matched design to minimise demographic confounding, the application of validated screening instruments, and a sample that reflects the ethnic composition of the UK population.

Limitations include the cross-sectional design, which precludes causal inference. We can there not infer whether there is a reverse-causality at play in which being unemployed increases the risk for mental health issues, which in turn could influence RAADS and ASRS scores. The sample was recruited via online platforms, which may over-represent digitally literate English-speaking UK adults and may not generalise to the most marginalised NEET populations. Self-report of physical and mental health conditions may introduce recall and reporting bias. Screening instruments are not diagnostic; screening-positive status reflects elevated trait burden rather than confirmed diagnosis. Future longitudinal studies should examine whether targeted pre-diagnosis support moderates NEET trajectories in those with elevated neurodivergent traits.

NEET populations are difficult to sample accurately from any online panel. The most disadvantaged NEET individuals may be missed because they are less likely to: (a) have stable internet access, (b) sign up for research platforms and (b) complete online surveys regularly. At the same time, people who are unemployed or economically inactive may have greater availability for paid online studies.

## Conclusion

These findings suggest that neurodivergent trait screening in NEET support services could identify individuals who would benefit from educational and occupational accommodations before formal diagnosis is secured. Employment advisors, Jobcentre Plus practitioners, and education providers should be trained to recognise neurodivergent presentations and to implement reasonable adjustments proactively. In line with the accessibility patterns shown here, policy and practice should prioritise adjustments that enhance sensory control, support condition management, and foster environments where neurodivergent adults can express themselves authentically, as these are the domains that NEET and EET participants with likely neurodivergence themselves identify as most consequential. Addressing the intersecting burden of physical health difficulties alongside neurodivergent needs, rather than treating these as separate service domains, is necessary to reduce NEET risk in this population. Reduction of NHS diagnostic waiting times and development of pre-diagnostic support pathways represent important systemic levers for improving employment and educational outcomes, not only for neurodivergent adults, but for the benefit of wider society.

## Author contributions

JE conceived this study. JE, LQ, and ER planned and designed this study. JE and LQ obtained funding. LQ collected data, conducted data analyses, interpreted and visualized data, and drafted the manuscript. BJ and EJ supported the first draft. UMS, CD, JG, JE, ER, and HC edited the manuscript. All authors approve the final version.

## Data availability

Data will be made available upon publication (link will be provided).

